# Chansu Injection Improves the Respiratory Function of Severe COVID-19 Patients

**DOI:** 10.1101/2020.05.20.20107607

**Authors:** Fen Hu, Jiao Chen, Hao Chen, Jin Zhu, Chen Wang, Haibin Ni, Jianming Cheng, Peng Cao, Xingxing Hu

## Abstract

Chansu and its major active constituent of bufalin have been reported to have broad-spectrum antiviral effects. This study aims to assess the efficacy of Chansu injection in treating patients with severe COVID-19. The patients diagnosed as severe or critical COVID-19 in The First People’s Hospital of Jiangxia District, Wuhan, China from February 5 to March 5, 2020 were randomly allocated in a 1:1 ratio to receive general treatment plus Chansu injection or only general treatment as the control group. The treatment course was 7 days. The changes of PaO_2_/FiO_2_ and ROX index indicating respiratory function, the white blood cell (WBC) count, peripheral blood mononuclear lymphocyte (PBML) count, respiratory support step-down time (RSST), and safety indicators for the 7th day were retrospectively analyzed. After 7 days treatment, the oxygenation index was improved in 20 of 21 patients (95.2%) in the treatment group, as compared with 13 of 19 patients (68.4%) in the control group. The PaO_2_/FiO_2_ and ROX indices in the treatment group (mean, 226.27±67.35 and 14.01±3.99 respectively) were significantly higher than the control group (mean, 143.23±51.29 and 9.64±5.54 respectively). The RSST was 1 day shorter in the treatment group than the control group. Multivariate regression analysis suggested that Chansu injection contributed the most to the outcome of PaO_2_/FiO_2_. No obvious adverse effects were observed. Preliminary data showed that Chansu injection had apparent efficacy in treating patients with severe COVID-19.

**Significance Statement:** Patients with severe COVID-19 usually have dyspnea and/or hypoxemia one week after the onset of symptoms, and can quickly progress to acute respiratory distress syndrome, septic shock, metabolic acidosis that is difficult to correct, coagulation dysfunction and multiple organs failure. Chansu injection, a widely used drug in clinical cancer and chronic hepatitis B therapy in China, can improve the respiratory function and shorten the respiratory support step-down time with 1 day in patients with severe COVID-19. These preliminary findings raise the possibility that Chansu injection may be helpful in the treatment of patients with severe COVID-19.

## Main Text

### Introduction

The current pandemic of SARS-CoV-2 is seriously threatening human health and socioeconomic development globally, which has plagued about 4,200,000 people and caused over 280,000 deaths by May 11, 2020. The disease of COVID-19 caused by SARS-CoV-2 is characterized by fever, dry cough, and fatigue. Severe patients usually have dyspnea and/or hypoxemia one week after the onset of symptoms, and can quickly progress to acute respiratory distress syndrome, septic shock, metabolic acidosis that is difficult to correct, coagulation dysfunction and multiple organs failure [1–4]. In severely affected areas such as Italy, the overall mortality rate is more than 10% according to the national official bulletin currently, and the mortality rate of critically severe patients is much higher. Therefore, reducing the mortality of severe or critically severe patients is very important. At present, no approved targeted therapeutics is available for COVID-19 and the treatment recommendation is also largely empirical. The Lopinavi-Ritonavir combination has been proved to have little effects on severe COVID-19 patients in clinical trials [5]. Based on China’s anti-epidemic experience during the outbreak in Wuhan, traditional Chinese medicine might play an important role in the treatment of COVID-19.

Chansu (Cinobufacini), aqueous extracts from the skin and parotid venom glands of the toad Bufo bufo gargarizans Cantor has been widely used in China as an cardiotonic, anodyne, antimicrobial, antineoplastic, and local anesthetic agent for thousands of years [6,7]. In the recent years, Chansu (in dosage forms of injection, capsules, oral solution and tablets) has been approved by the Chinese State Food and Drug Administration (SFDA) and widely used in clinical cancer and chronic hepatitis B therapy in China [8,9]. Previous studies found that, bufalin, the major active constituent of Chansu can inhibit infection of cells with murine hepatitis virus (MHV), feline infectious peritonitis virus (FIPV), Middle East respiratory syndrome (MERS-CoV), and vesicular stomatitis virus (VSV) by targeting the ATP1A1-mediated Src signaling pathway [10], which also plays a critical role in Ebola and respiratory syncytial virus (RSV) entry into host cells [11,12]. Based on these findings, Chansu injection was empirically applied to patients with severe COVID-19. Controlled studies are difficult to perform in an epidemic of such a life threatening condition. We present the results from a randomized, preliminary clinical study that evaluated the effect of adding Chansu injection to a general treatment in patients with severe COVID-19.

## Results

### Patient Characteristics

This study preliminary evaluated the efficacy and safety of Chansu injection for severe COVID-19 patients infected by SARS-CoV-2 (trial registration: ChiCTR2000030704). A total of 50 patients were assessed for eligibility and randomly allocated into general treatment (the control group) or general treatment plus Chansu injection group (the treatment group) in a 1:1 ratio. Ten patients were excluded from the final analysis, including 3 patients in the treatment group and 6 patients in the control group transferring to another hospital for further treatment, and 1 patient in the treatment group failed to re-examination the indicators on schedule. The remaining 40 patients including 21 in the treatment group and 19 in the control group were included for statistical analysis (Figure 1).

**Figure 1.**
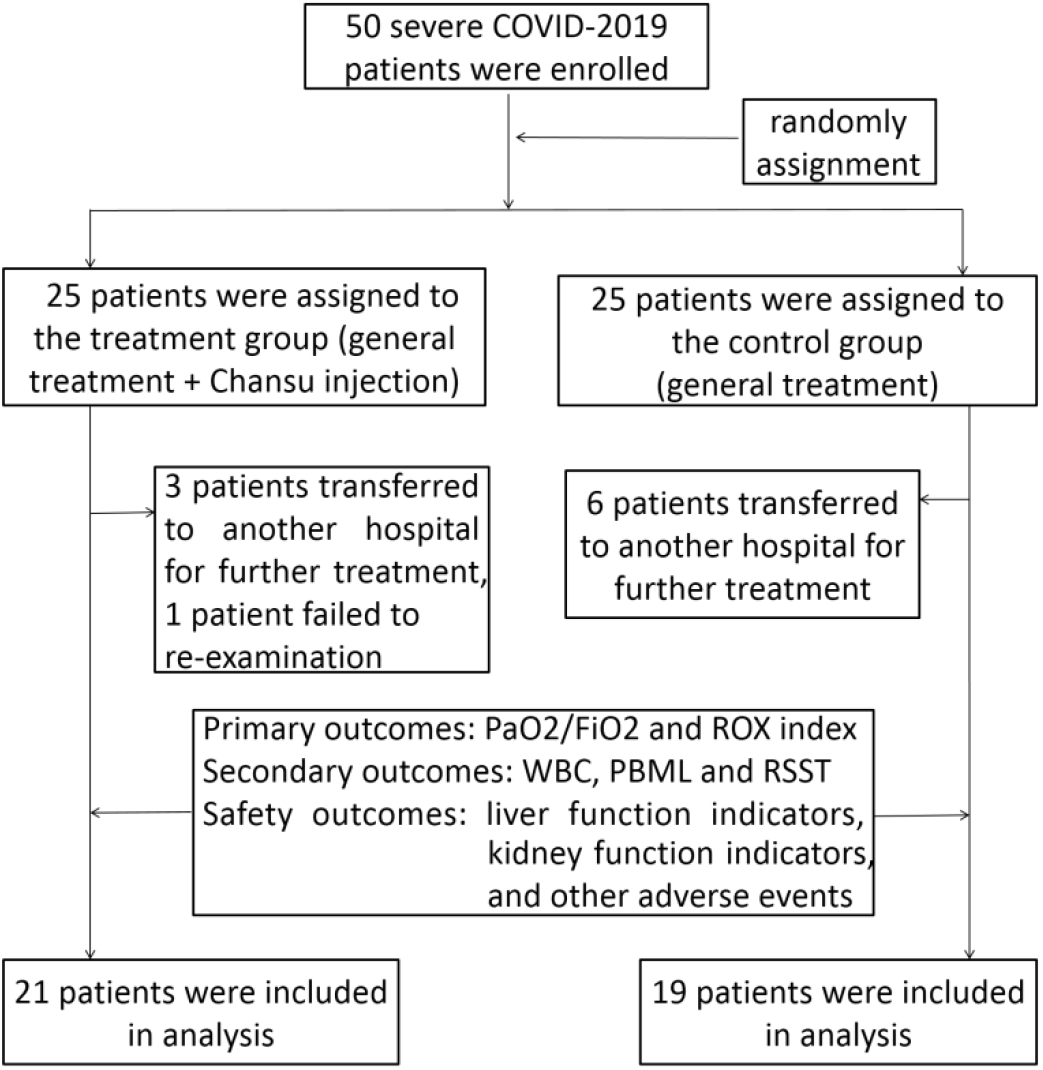
Randomization and Treatment Assignment.

Of the 40 participants, 24 (60%) were male. The median age was 61.5 years old (interquartile range [IQR], 51.0 to 71.0 years) with most patients (82.5%) >50 years old. At least one third of the patients have one or more basic diseases. The median time from onset to treatment was 6 days (IQR, 4.0 to 7.8 days). 36 (90%) patients had a body temperature >37 centigrade at enrollment. The mean SpO_2_ was 88.73%. No significant differences were found between the treatment group and control group in gender, age, BMI, time from onset to treatment, basic diseases and vital signs at enrollment (*P* > 0.05, Table 1).

**Table 1.** Baseline characteristics of the participants.

|  | **All patients**<br>**(n=40)** | **Treatment**<br>**Group (n=21)** | **Control**<br>**Group (n=19)** | **P**<br>**Value** |
| --- | --- | --- | --- | --- |
| **Gender (Male, n, %)** | 24 (60) | 11 (52.38) | 13 (68.42) | 0.301 |
| **Age (yr), median (IQR)** | 61.5 (51.0-71.0) | 58.0 (52.5-66.0) | 70.0 (51.0-77.0) | 0.283 |
| **BMI (kg/m<sup>2</sup>), median (IQR)** | 26 (23.6-27.2) | 26.4 (24.6-27.3) | 25.2 (22.3-26.9) | 0.391 |
| **Time from onset to treatment**<br>**(day), median (IQR)** | 6.0 (4.0-7.8) | 7.0 (4.0-7.5) | 5.0 (3.0-8.0) | 0.685 |
| **Basic Diseases (n, %)** |  |  |  |  |
| High Blood Pressure | 12 (30.00) | 5 (23.81) | 7 (36.84) | 0.369 |
| Diabetes | 9 (22.50) | 5 (23.81) | 4 (21.05) | 0.835 |
| Chronic Obstructive Pulmonary Disease | 1 (2.50) | 0 | 1 (5.26) | 0.475 |
| Coronary Heart Disease | 2 (5.00) | 0 | 2 (10.53) | 0.127 |
| Chronic Cardiac Insufficiency | 1 (2.50) | 0 | 1 (5.26) | 0.475 |
| **Vital signs at enrollment<sup>^</sup>** |  |  |  |  |
| Temperature (°C) | 38.01±0.93 | 38.18±0.94 | 37.82±0.90 | 0.230 |
| RR (bpm) | 24.95±4.30 | 25.48±4.59 | 24.37±4.00 | 0.423 |
| HR (bpm) | 94.88±19.62 | 99.67±13.54 | 89.58±23.96 | 0.105 |
| SpO<sub>2</sub> (%) | 88.73±6.12 | 88.05±7.65 | 89.47±23.96 | 0.469 |

BMI: body mass index, RR: respiratory rate, HR: heart rate, SpO<sub>2</sub>: pulse oxygen saturation
P value: statistical significance between the treatment group and control group
<sup>^</sup>: Plus-minus values are means ±SD

### Outcomes

Compared with CT imaging characteristics and various biochemical indicators, the patients’ respiratory function indicators, such as PaO_2_/FiO_2_, SpO_2_ and RR related ROX index are more direct indicators of disease outcome. Therefore, PaO_2_/FiO_2_ and ROX index were used as the primary outcomes in this study. No significant difference was found in the initial PaO_2_/FiO_2_ and ROX index on Day 1 between the treatment group and control group (*P* = 0.118, 95% CI, -6.46 to 55.18 for PaO_2_/FiO_2_; *P* = 0.716, 95% CI, -1.89 to 2.73 for ROX) (Table 2). After 7 days treatment, patients receiving general treatment plus Chansu injection (20 mL/day) improved significantly in both PaO_2_/FiO_2_ and ROX index (*P* < 0.001, 95% CI, -111.30 to -35.90 for PaO_2_/FiO_2_; *P* < 0.001, 95% CI, -7.56 to -2.94 for ROX), while no significant differences were found in the control group receiving only general treatment (*P* > 0.05). The PaO_2_/FiO_2_ and ROX index were improved in 20 patients (95.2%) in the treatment group, as compared with 13 (68.4%) and 14 (73.7%) patients in the control group (Table 2). These results indicated that most severe COVID-19 patients benefited from Chansu injection, while general treatment with empirical antiviral therapy such as paramivir, arbidol and interferon α, standard glucocorticoid therapy or other symptom relievers and oxygen therapy had little effect on severe COVID-19 patients.

COVID-19 patients usually have normal or decreased WBC and decreased PBML. For the secondary outcomes of WBC and PBML, no obvious improvement was observed after 7 days treatment in both groups (*P* > 0.05). RSST is the time needed for the patients to transform from advanced respiratory support to low respiratory support. The shorter RSST is associated with the faster prognosis of patients’ respiratory function. The median of RSST was 5.0 (IQR, 4.0 to 6.0) days in the treatment group, 1 day shorter than that in the control group (median: 6.0 days, IQR: 5.0 to 8.0 days) (Table 2).

**Table 2.**
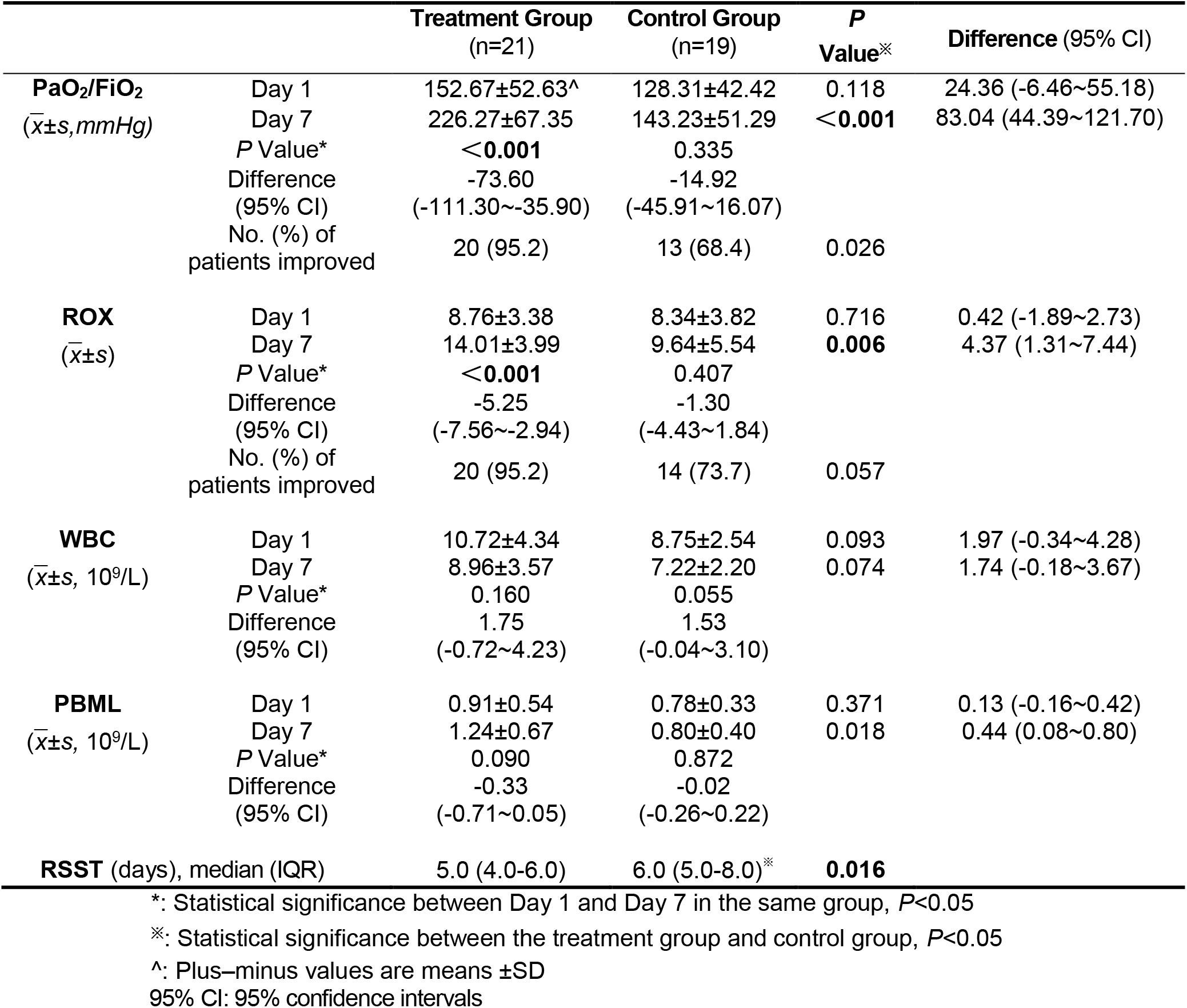
Comparison of the outcomes between the treatment group and control group.

|  |  | Treatment Group<br>(n=21) | Control Group<br>(n=19) | P<br>Value* | Difference (95% CI) |
| --- | --- | --- | --- | --- | --- |
| **PaO<sub>2</sub>/FiO<sub>2</sub>**<br>($\bar{x} \pm s, mmHg$) | Day 1 | 152.67±52.63<sup>^</sup> | 128.31±42.42 | 0.118 | 24.36 (-6.46~55.18) |
| | Day 7 | 226.27±67.35 | 143.23±51.29 | **<0.001** | 83.04 (44.39~121.70) |
|  | P Value* | **<0.001** | 0.335 |  |  |
|  | Difference<br>(95% CI) | -73.60<br>(-111.30~-35.90) | -14.92<br>(-45.91~16.07) |  |  |
|  | No. (%) of<br>patients improved | 20 (95.2) | 13 (68.4) | 0.026 |  |
| **ROX**<br>($\bar{x} \pm s$) | Day 1 | 8.76±3.38 | 8.34±3.82 | 0.716 | 0.42 (-1.89~2.73) |
| | Day 7 | 14.01±3.99 | 9.64±5.54 | **0.006** | 4.37 (1.31~7.44) |
|  | P Value* | **<0.001** | 0.407 |  |  |
|  | Difference<br>(95% CI) | -5.25<br>(-7.56~-2.94) | -1.30<br>(-4.43~1.84) |  |  |
|  | No. (%) of<br>patients improved | 20 (95.2) | 14 (73.7) | 0.057 |  |
| **WBC**<br>($\bar{x} \pm s, 10^9/L$) | Day 1 | 10.72±4.34 | 8.75±2.54 | 0.093 | 1.97 (-0.34~4.28) |
| | Day 7 | 8.96±3.57 | 7.22±2.20 | 0.074 | 1.74 (-0.18~3.67) |
|  | P Value* | 0.160 | 0.055 |  |  |
|  | Difference<br>(95% CI) | 1.75<br>(-0.72~4.23) | 1.53<br>(-0.04~3.10) |  |  |
| **PBML**<br>($\bar{x} \pm s, 10^9/L$) | Day 1 | 0.91±0.54 | 0.78±0.33 | 0.371 | 0.13 (-0.16~0.42) |
| | Day 7 | 1.24±0.67 | 0.80±0.40 | 0.018 | 0.44 (0.08~0.80) |
|  | P Value* | 0.090 | 0.872 |  |  |
|  | Difference<br>(95% CI) | -0.33<br>(-0.71~0.05) | -0.02<br>(-0.26~0.22) |  |  |
| **RSST (days), median (IQR)** |  | 5.0 (4.0-6.0) | 6.0 (5.0-8.0)* | **0.016** |  |

\*: Statistical significance between Day 1 and Day 7 in the same group, $P < 0.05$
※: Statistical significance between the treatment group and control group, $P < 0.05$
<sup>^</sup>: Plus-minus values are means ± SD
95% CI: 95% confidence intervals

### Chansu injection contributes the most to the primary outcome of PaO_2_/FiO_2_

Univariate linear regression analyses were performed on the patients’ baseline characteristics, PaO_2_/FiO_2_, ROX index, WBC and PBML on Day 1, and treatment interventions to screen out variables with regression coefficients statistically significant. Four characteristics: the PaO_2_/FiO_2_ and PBML on Day 1, the initial respiratory support mode and Chansu injection affected PaO_2_/FiO_2_ on Day 7 statistically (*P* < 0.05). To determine the main variables affecting the primary outcome of PaO_2_/FiO_2_ on Day 7, multivariate regression analysis was applied. The PaO_2_/FiO_2_ on Day 1, the initial respiratory support mode and Chansu injection were further confirmed that could affect the primary outcome of PaO_2_/FiO_2_ on Day 7 statistically (*P* < 0.05). Among them, Chansu injection contributed to the most to the regression model (Beta = 0.486) (Table 3).

**Table 3.** Multivariate regression analysis to determine the most contributing factor to the primary outcome of PaO_2_/FiO_2_

| Model | Unstandardized Coefficients |  | Standardized Coefficients | t | *P* Value | Collinearity Statistics |  |
| --- | --- | --- | --- | --- | --- | --- | --- |
|  | B | Std.Error | Beta |  |  | Tolerance | VIF |
| Constant | 113.317 | 32.888 |  | 3.446 | .001 |  |  |
| PaO<sub>2</sub>/FiO<sub>2</sub> (Day 1) | .513 | .170 | .346 | 3.019 | .005 | .744 | 1.344 |
| Initial respiratory support mode | -19.093 | 6.666 | -.314 | -2.864 | .007 | .812 | 1.231 |
| Chansu injection | 69.969 | 15.321 | .486 | 4.567 | .000 | .863 | 1.159 |
| PBML (Day 1) | 9.137 | 13.525 | .676 | .676 | .504 | .843 | 1.186 |

### Safety

No obvious adverse events such as systemic or local rash, gastrointestinal symptoms, abdominal pain and diarrhea or new arrhythmia occurred during treatment with Chansu injection. No significant difference in other safety outcomes including liver function indicators (AST, ALT, TB), kidney function indicators (Cr), CK-MB and PLT was found before and after treatment with Chansu injection (P >0.05). No significant difference were observed between the treatment group and control group (Supplementary Table S1). This indicated that the dose of Chansu injection used in this study is safe for severe COVID-19 patients.

## Discussion

At present, no approved targeted therapeutics is available for COVID-19 and the treatment recommendations are largely empirical. Chansu is a kind of traditional Chinese medicine extracted from the skin and parotid glands of the toad, which has been approved by SFDA for cancer and antiviral therapy in China. In this study, we evaluated the effect of Chansu plus general treatment in patients with severe COVID-19 in a randomized, preliminary clinical trial.

This randomized trial found that, compared with general treatment alone, Chansu injection added to general treatment significantly improved the primary outcomes of PaO_2_/FiO_2_ and ROX index in patients with severe COVID-19 after 7 days treatment. General treatment with empirical antiviral therapy such as paramivir, arbidol and interferon α, standard glucocorticoid therapy or other symptom relievers and oxygen therapy had little effect on severe COVID-19 patients. From clinical experience, we found that patients’ respiratory function status, such as SpO_2_, PaO_2_/FiO_2_, and ROX index are more direct indicators of an improvement of this disease. In addition, the median RSST needed for the patients in the treatment group is 5.0 days, about one day shorter than the control group. Multivariate regression analysis suggested that, among various variables, Chansu injection contributed the most to the primary outcomes of the patients in the treatment group.

The main active ingredients of Chansu include bufalin, resibufogenin and cinobufagin, etc. Researchers have found that Chansu has various activities including anti-inflammatory, anti-infection and immunomodulatory [13–15]. Recent studies reported that Chansu or its active ingredient bufalin shows obvious anti-HBV activities and can inhibit infection of cells with MERS-CoV, Ebola, RSV, MHV, FIPV, and VSV [8–10]. The mechanism of Chansu inhibiting coronavirus entry into host cells might be related to ATP1A1-mediated Src signaling pathway [8–10]. Recently, researchers identified ATPases as one of the target of SARS-CoV-2 by interacting with Nsp6 using affinity-purification mass spectrometry method [16]. However, how the coronavirus interacts with ATPases needs further investigation.

The side effects caused by Chansu are mainly related to its cardiac glycoside property [17]. In this clinical trial, no obvious adverse events occurred in the patients receiving Chansu injection. Our preliminary results suggests that the dose of Chansu injection (20 mL/day, containing about 60 μg dry extract of Chansu) used in this study is safe and effective in treating patients with severe COVID-19. Such results are promising and give an alternative strategy to fight the emerging SARS-CoV-2 infection in real-time. Given the urgent need for effective drugs against SARS-CoV-2 in the current pandemic condition, we recommend that severe COVID-19 patients be treated with Chansu to cure the disease and improve their respiratory function.

This clinical trial conducted in such an emergency has several limitations. Firstly, the trial was not blinded, which might influence clinical decision-making. Secondly, because of limited conditions, the effect of virologic clearance was not tested after 7 days treatment. We can only use the outcomes of respiratory function such as PaO_2_/FiO_2_ and ROX index to evaluate the efficacy of Chansu injection. Thirdly, some patients had a history of out-of-hospital treatment, leading to the inconsistency of disease course. Interventions not being able to carried out at the same time after the patients transformed from mild symptoms to severe symptoms may lead to biased results. Other limitations include a small sample size and limited long-term outcome follow-up.

In conclusion, we found that Chansu has apparent efficacy in treating severe COVID-19 in this preliminary study. It can improve the patients’ respiratory function significantly. These promising early data provide a reference for the treatment of severe COVID-19.

## Materials and Methods

### Study Design and Participants

A prospective randomized controlled study was conducted. From February 5, 2020 to March 5, 2020, a total of 50 patients with severe or critical severe COVID-19 admitted to the department of Respiratory and Critical Care Medicine of The First People’s Hospital of Jiangxia District, Wuhan, China were enrolled in the trial after approval by the ethics committees. All patients were confirmed COVID-19 by a reverse-transcriptase-polymerase-chain-reaction (RT-PCR) assay or clinical diagnosis according to the Guidelines for the Prevention, Diagnosis, and Treatment of Pneumonia Caused by COVID-19 (version 5) issued by the National Health Commission of the People’s Republic of China. Severe or critical severe patients meet at least one of the following diagnostic criteria: respiratory rate (RR) ≥30 bpm in calm state, pulse oxygen saturation (SpO_2_) ≤93% while breathing ambient air in resting state, oxygenation index (a ratio of the arterial partial pressure of oxygen to the fraction of inspiration oxygen, PaO_2_/FiO_2_) ≤300 mmHg or respiratory failure (PaO_2_ ≤60 mmHg with or without carbon dioxide retention under standard conditions). Main exclusion criteria included age <7 years old, pregnant female, history of arrhythmia, chronic respiratory failure caused by other diseases, such as heart failure, thoracic deformity, structural lung disease, etc., hemodynamic instability, severe immunodeficiency or using immunosuppressants recently, allergies, estimated survival time <3 days, withdrawal from research or return visits. Written informed consent was obtained from all patients or from the patient’s legal representative. The trial was conducted in accordance with the principles of the Declaration of Helsinki and the Good Clinical Practice guidelines of the International Conference on Harmonisation.

### Randomization and trial procedures

The enrolled patients were randomly assigned in a 1:1 ratio using a random-numbers table to receive general treatment (the control group) or general treatment plus Chansu injection (20 mL/day, containing about 60 μg dry extract of Chansu. Jiangsu Pujin Pharmaceutical Co., Ltd., Batch number: 191002, SFDA approval number: Z32020694). General treatment includes, as necessary, empirical antiviral therapy with peramivir, arbidol and interferon α, nutritional support, supplemental oxygen and standard glucocorticoid therapy, etc (Supplementary Table S2). The treatment group was given Chansu injection (20 mL formulated into 250 mL 0.9% physiological saline) intravenously at the speed of 125 mL/h every day, while the control group was given 250 mL 0.9% physiological saline. The study interventions continued for 7 days until intolerable adverse events, withdrawal of consent or transferring to another hospital for further treatment.

### Baseline and Outcomes

Baseline characteristics include gender, age, body mass index (BMI), time from onset to treatment, basic diseases, and vital signs including body temperature, RR, heart rate, and SpO_2_ etc. at the time of enrollment.

Primary outcomes include PaO_2_/FiO_2_ and ROX index (ROX=SpO_2_/(FiO_2_*RR)). Secondary outcomes include white blood cell (WBC) count, peripheral blood mononuclear lymphocyte (PBML) count and respiratory support step-down time (RSST). Four respiratory supports of non-invasive ventilation, high-flow oxygen therapy, oxygen mask, and oxygen support with nasal duct gradually weakens for the patients. Therefore, RSST is defined as the transition time from advanced respiratory support to low respiratory support after enrollment, or the time required for FiO_2_ to decrease by 50%. Successful respiratory support transition is considered if SpO_2_ remained ≥94% and the increase of RR did not exceed 20%.

Safety outcomes include liver function indicators (AST, ALT, TB), kidney function indicators (Cr), CK-MB, PLT and adverse events such as systemic or local rash that cannot be explained by other reasons, gastrointestinal symptoms including nausea, vomiting, abdominal pain and diarrhea, new arrhythmia, etc. Adverse events were assessed according to the National Cancer Institute Common Terminology Criteria for Adverse Events, version 4.0.

### Statistical Analysis

All data analyses were performed in SPSS 22.0. Continuous variables were summarized as medians and ranges or arithmetic means with standard deviations. Student’s t test was used for assessing statistical significance of continuous variables between the control and treatment groups. Categorical variables were summarized as percentages (%) and the χ^2^ test was used for assessing their statistical significance between them. Multiple linear regression analysis was performed to evaluate the effect of the variables on the primary outcome by adjustment of imbalance for important baseline variables because of withdraw of some patients. Univariate linear regression analysis was performed to screen out variables with regression coefficients statistically significant (*P* < 0.05), which were further included in the multivariate linear regression to evaluate their effects on the primary outcome.

## Data Availability

The data used to support the findings of this study are available from the corresponding author upon request.

## Acknowledgments

This work was supported by a project funded by the Priority Academic Program Development of Jiangsu Higher Education Institutions (Integration of Chinese and Western Medicine).

## References

[1] N. Zhu, D. Zhang, W. Wang, et al. A Novel Coronavirus from Patients with Pneumonia in China. N. Engl. J. Med. DOI: 10.1056/NEJMoa2001017 (2019).

[2] C. Huang, Y. Wang, X. Li, et al. Clinical features of patients infected with 2019 novel coronavirus in Wuhan, China. Lancet 395: 497–506 (2020).

[3] Z. Wu, J.-M. McGoogan. Characteristics of and important lessonsfrom the coronavirus disease 2019 (COVID-19) outbreak in China: summary of a report of 72 314 cases from the Chinese Center for Disease Control and Prevention. JAMA. doi: 10.1001/jama.2020.2648 (2020).

[4] N. Chen, M. Zhou, X. Dong, et al. Epidemiological and clinical characteristics of 99 cases of 2019 novel coronavirus pneumonia in Wuhan, China: a descriptive study. Lancet 395: 507–13 (2020).

[5] B. Cao, Y. Wang, D. Wen, et al. A Trial of Lopinavir–Ritonavir in Adults Hospitalized with Severe Covid-19. N. Engl. J. Med. DOI: 10.1056/NEJMoa2001282 (2020).

[6] F. Qi, A. Li, L. Zhao, et al. Cinobufacini, an aqueous extract from Bufo bufo gargarizans Cantor, induces apoptosis through a mitochondria-mediated pathway in human hepatocellular carcinoma cells. J. Ethnopharmacol. 128:654–661 (2010).

[7] F. Qi, A. Li, Y. Inagaki, et al. Antitumor activity of extracts and compounds from the skin of the toad Bufo bufo gargarizans Cantor. Int. Immunopharmacol. 11:342–349 (2011).

[8] F. Qi, Z. Wang, P. Cai, et al. Traditional Chinese medicine and related active compounds: A review of their role on hepatitis B virus infection. Drug Discov. Ther. 7(6):212–224 (2013).

[9] J. Qi, C.-K. Tan, S.-M. Hashimi, et al. Toad Glandular Secretions and Skin Extractions as Anti-Inflammatory and Anticancer Agents. Evid. Based Complement. Alternat. Med. 2014:312684 (2014).

[10] C. Burkard, M.-H. Verheije, B.-L. Haagmans, et al. ATP1A1-Mediated Src Signaling Inhibits Coronavirus Entry into Host Cells. J. Virol. 89(8):4434–48 (2015).

[11] I. García-Dorival, W. Wu, S. Dowall, et al. Elucidation of the Ebola Virus VP24 Cellular Interactome and Disruption of Virus Biology through Targeted Inhibition of Host-Cell Protein Function. J. Proteome Res. 13(11):5120–35 (2014).

[12] M. Lingemann, T. McCarty, X. Liu, et al. The alpha-1 subunit of the Na+, K+-ATPase (ATP1A1) is required for macropinocytic entry of respiratory syncytial virus (RSV) in human respiratory epithelial cells. PLoS Pathog. 15(8):e1007963 (2019).

[13] J.-Y. Wang, L. Chen, Z. Zheng, et al. Cinobufocini inhibits NF-*κ*B and COX-2 activation induced by TNF-*α*in lung adenocarcinoma cells. Oncol. Rep. 27(5):1619–1624 (2012).

[14] G.-A. Cunha Filho, C.-A. Schwartz, I.-S. Resck, et al. Antimicrobial activity of the bufadienolides marinobufagin and telocinobufagin isolated as major components from skin secretion of the toad Bufo rubescens. Toxicon 45(6):777–782 (2005).

[15] X.-L. Wang, G.-H. Zhao, J. Zhang, et al. Immunomodulatory effects of cinobufagin isolated from ChanSu on activation and cytokines secretion of immunocyte in vitro. J. Asian Nat. Prod. Res. 13(5):383–392 (2011).

[16] D.-E. Gordon, G.-M. Jang, M. Bouhaddou, et al. A SARS-CoV-2 protein interaction map reveals targets for drug repurposing. Nature doi: 10.1038/s41586-020-2286-9 (2020).

[17] M. GowdaR, R.-A. Cohen, I.-A. Khan. Toad venom poisoning: resemblance to digoxin toxicity and therapeutic implications. Heart 89(4):e14 (2003).

